# Combined therapy of prednisone and mTOR inhibitor sirolimus for treating retroperitoneal fibrosis

**DOI:** 10.1101/2022.09.05.22279516

**Authors:** Hui Gao, Shibo Liu, Yuanbang Mai, Yuying Wang, Xue-wu Zhang, Shufen Zheng, Chenghua Luo, Cuiping Pan

## Abstract

Retroperitoneal fibrosis (RPF) is a rare autoimmune disease with fibrous tissue growth and inflammation in retroperitoneum, whose development could encase surrounding organs and lead to severe conditions. Its current treatments involve long-term uptake of glucocorticoids (e.g., prednisone) for controlling inflammation; however, side effects are common, triggering search for replacement therapies. Here, we surveyed gene-disease databases and discovered that mTOR displayed significant changes in RPF, which we confirmed by immunohistological staining. Next, we inferred from drug-gene databases that mTOR inhibitor compound sirolimus could affect most biological pathways in RPF. We then designed a combined therapy in which a gradual reduction of prednisone was prescribed with a long-term, stable dosage of sirolimus. We implemented a single-arm clinical trial in RPF patients and assessed the treatment effects at three timepoints (0, 12 weeks and 48 weeks of treatment). By assessing fibrous tissue mass by computed tomography, inflammation markers and kidney functions by lab tests, immune cell types and abundances by flow cytometry, and plasma inflammation-related proteins by Olink proteomics, we revealed that our combined therapy resulted in significant fibrosis remission and an overall regression of the immune system towards healthy states. In addition, no obvious side effects were observed. We concluded that this new therapy had the potential to replace long-term steroid monotherapy for treating RPF.

## Introduction

Retroperitoneal fibrosis (RPF), also called Ormond’s disease, periureteritis fibrosa, periureteritis plastica, chronic periureteritis, and fibrous retroperitonitis, is a rare autoimmune disease characterized by the presence of inflammatory and fibrous tissue in the retroperitoneum. Its prevalence is estimated at 1.4 cases/100,000 inhabitants and incidence at 0.1-1.3 cases/100,000 persons per year [1,2]. Typical symptoms of RPF include pain in the lower back and abdomen, weight loss, fever, nausea and anemia. In severe cases, the growth of the fibrous tissue can encase important organs, such as the abdominal aorta, iliac arteries and ureters, causing inflammatory abdominal aortic aneurysm and ureteral obstruction [3]. Importantly, ureteral obstruction occurs in about 60-80% of cases and often causes chronic kidney disease, end-stage renal diseases and kidney atrophy [1].

RPF may be primary (idiopathic) or secondary. Idiopathic RPF develops with no known etiology and accounts for more than 70 percent of the cases [1,2]. Some of the RPF patients have elevated immunoglobulin G4 (IgG4) levels, multi-organ inflammation and fibrosis, hence belonging to IgG4-related diseases (IgG4-RD), whereas others have normal IgG4 levels [4]. Secondary RPF often results from infections, malignancy, drugs, retroperitoneal hemorrhage or various other disorders. Risk factors of RPF include genetic predisposition, e.g., the HLA-DRB1*03 allele [5], and environmental exposures including asbestos and smoking [6].

Biological mechanisms of RPF have begun to emerge [1]. It has been shown that recruitment of CD4^+^ T cells in the lesions would release cytokines, such as interleukin (IL)-6 and IL-21, and attract B cells to form germinal centers (GCs) in which cytokines including IL-4, IL-6, IL-10, IL-13 and eotaxin-1 are further produced. Additional immune cells such as eosinophils and mast cells are found in the microenvironment. With a complex interaction among the cells and cytokines, fibroblasts ultimately differentiate into myofibroblasts and produce massive amounts of collagen, forming the fibrous mass. Many of these reactions resemble other autoimmune conditions that involve fibrosis, such as systemic sclerosis (SSc) and interstitial lung disease (ILD) [7,8].

The current mainstay therapy for idiopathic RPF is synthetic glucocorticoids, e.g., prednisone, for suppressing inflammatory responses [9,10]. These are steroid hormones that mimic the endogenous hormone *cortisol* in both its structural and pharmacological features. Prednisone are small, lipophilic substances that can diffuse through the cell membrane and bind to the glucocorticoid receptors in cytoplasm, forming complexes. These complexes exert immunosuppressive effects in several ways [11]. First, they can bind to transcription factors and repress transcription of pro-inflammatory cytokines, such as IL-1, IL-2, IL-6, IL-8, tumor necrosis factor (TNF), interferon (IFN)-gamma, Cox-2, vascular endothelial growth factor (VEGF) and prostaglandins. Furthermore, the complexes can activate other transcription factors, leading to the production of anti-inflammatory cytokines, such as IL-10, nuclear factor kappa B (NF-κB) inhibitor, and lipocortin-1. In addition, high doses of glucocorticoids act via non-transcriptional mechanisms by binding to the glucocorticoid receptors in immune cells and therefore impair their signal transduction and immune responses. Other effects include decreasing the number of immune cells, reducing proliferation of fibroblasts, decreasing phagocytosis and antigen presentation by macrophages, inhibiting metalloproteinases collagenase and stromelysin, etc.

While glucocorticoids have been established in the last decade as a standard medication to treat idiopathic RPF, side effects of steroid hormones are often observed, especially when used for extended periods [12]. In clinical practices, glucocorticoids are prescribed at a dosage of 0.6-1 mg/kg/day for patients without contraindications, and then tapered to a minimal dosage (generally equivalent to less than 7.5 mg per day of prednisone) or stopped. However, reducing dosages triggers more frequent disease recurrence varying from 17.6 to 72% [1,13-16]. An effective and safe therapy has yet to be defined for RPF.

Here we leveraged drug target databases and identified *in silico* potential chemical compounds that could impact RPF. First, we discovered that the PI3K-mTOR-AKT pathway was highly activated in RPF tissues, and *sirolimus*, an mTOR inhibitor, appeared as a good treatment candidate as it targeted most of the biological pathways in RPF. Next, we designed a single-arm clinical trial of a combined therapy with prednisone and sirolimus, and assessed their effects on RPF patients in a 48-week follow-up study. Our results showed that this combined therapy effectively reduced fibrous mass and restored immune profiles in RPF patients, suggesting it has potential to replace the long-term monotherapy of steroid hormone for RPF.

## Results

### mTOR pathway was highly activated in RPF

Searching the human gene-disease databases MalaCards [17] and DisGenet [18], 14 genes in total appeared as relevant to RPF (Supplemental Table 1). The genes identified in MalaCards were enriched for cytotoxicity and kinase pathways, while the genes in DisGenet were enriched for cytokine and chemokine pathways (adjusted P < 0.05). Interestingly, only two genes overlapped, suggesting the two databases captured different biological aspects of the disease (Supplemental Fig 1). Next, we searched in GeneCards for RPF, which was an ensemble knowledgebase integrating genome, transcriptome, proteome, genetics, clinical and functional information from around 150 web sources [19]. The search resulted in around 500 genes, covering nearly all the enriched gene ontology (GO) terms and pathway terms (Kyoto Encyclopedia of Genes and Genomes, KEGG) by the previous MalaCards and DisGenet analyses, corroborating the diverse disease biology. For a broad understanding of the pathology of RPF, we pooled all the genes identified from the three databases and performed enrichment analysis collectively. A few key signaling pathways were discovered, including the mammalian target of rapamycin (mTOR) pathway and its associated phosphoinositide-3-kinase (PI3K)-Akt pathway (Figure 1A).

**Table 1.**
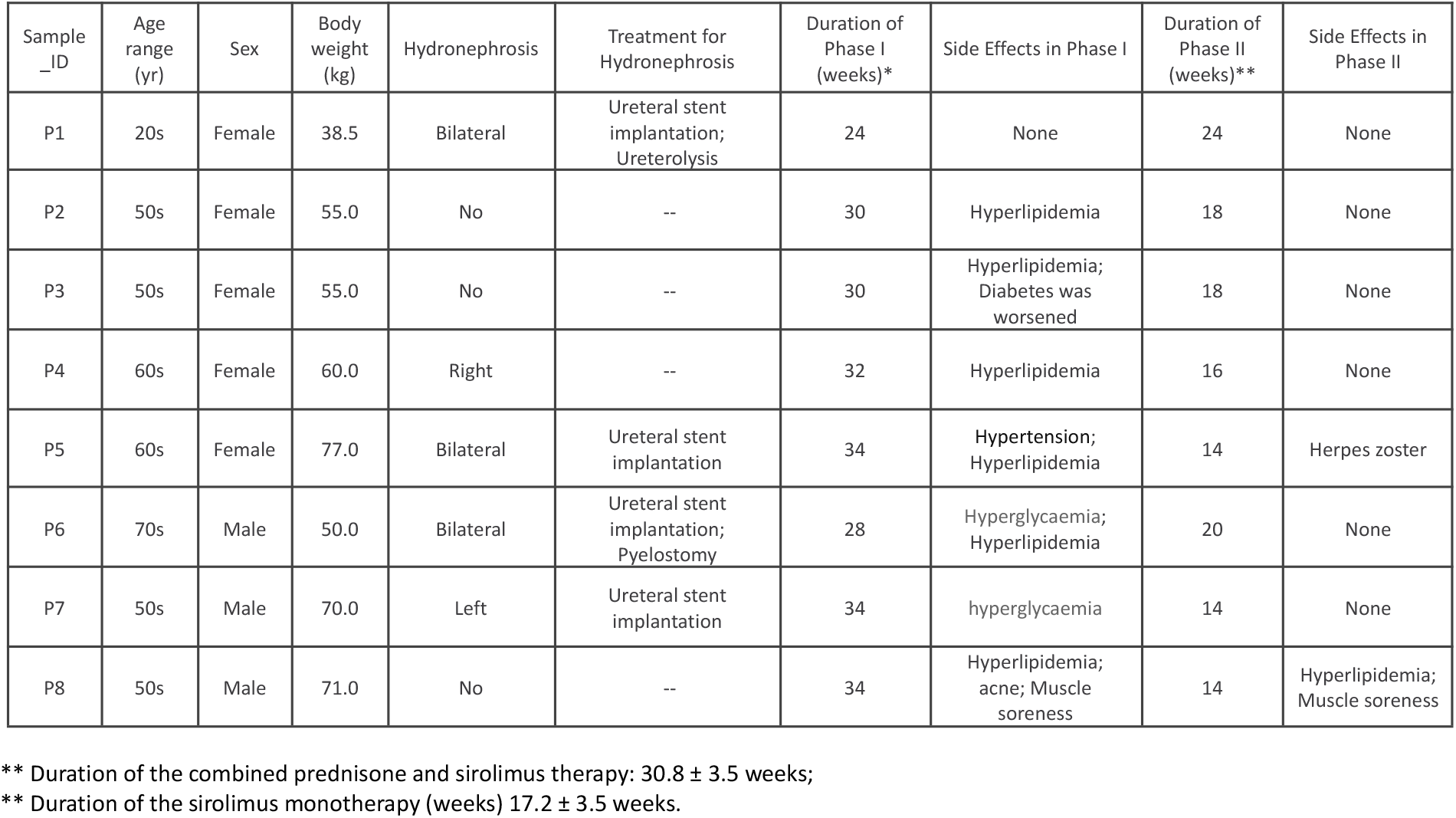
A summary of the RPF patients (n=8) and their treatments with combined therapy of prednisone and sirolimus.

**Figure 1.**
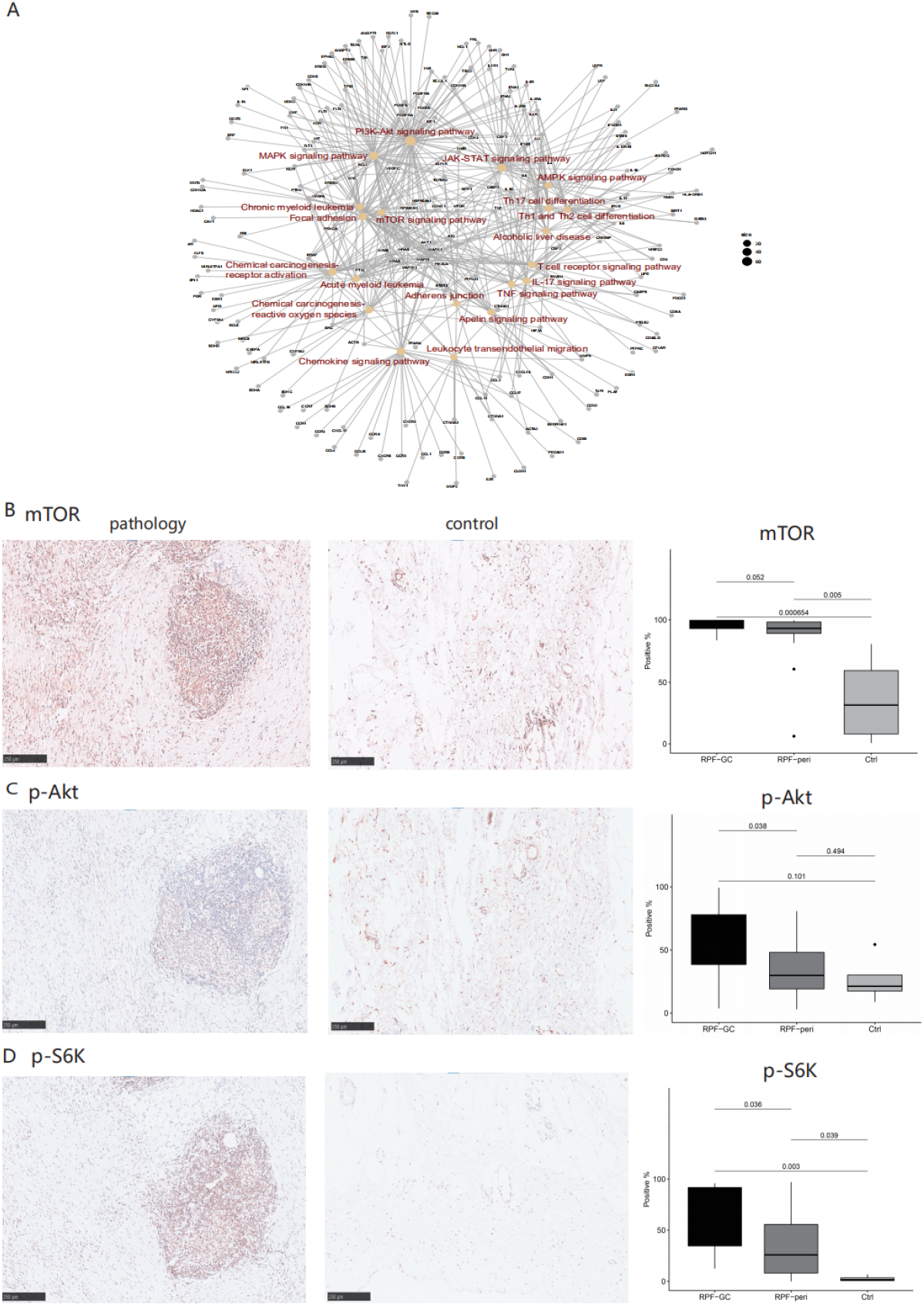
The PI3K-AKT-mTOR pathway was highly activated in RPF tissues. (A) Signaling pathway network enriched in genes related to RPF. (B-D) Immunohistological staining of mTOR and its downstream signaling proteins, phospho-Akt and phospho-S6K, with the boxplots on the right displaying the percentages of positive staining in 16 patients for RPF tissues (RPF-GC and RPF-peri) and and 4 controls (Ctrl). RPF-GC: retroperitoneal fibrosis geminal center; RPF-peri: retroperitoneal fibrosis peripheral; Ctrl: healthy control.

For validating their activities, we used antibodies to stained the key molecules mTOR and its two downstream signaling molecules, phospho-S6K (p-S6K) in mTOR-complex 1 and phospho-AKT (p-Akt) in mTOR-complex 2; in addition, we examined them at three locations of the fibrous tissues: the germinal centers (GC) where the fibrosis was most dense with a high infiltration of T and B cells, the peripheral that was proximal to the GCs, and normal controls (Figure 1B-1D). As normal controls were hard to obtain in healthy individuals, we chose the mesenteric root tissue biopsies derived from early-diagnosed colon cancer patients who were confirmed by pathological examination to have no lymphatic metastasis at the time of biopsy. We observed > 90% of cells stained for mTOR and p-S6K in the fibrous tissues, whether in the GCs or peripherals. p-AKT was also activated, albeit to a lesser degree. Therefore, we concluded that mTOR-complex 1 was highly activated in both the GC and peripheral of the fibrous tissues.

### The mTOR inhibitor Sirolimus affects most of the RPF pathways

Learning of the high mTOR activity in RPF, we analyzed *in silico* if the mTOR inhibitor compound, sirolimus, could potentially target RPF pathways. Sirolimus, also known as rapamycin, is an immunosuppressive drug that binds to the intracellular protein FKBP-12, forming a complex that targets the kinase mTOR, leading to inhibition of cytokine receptor-dependent signal transduction, deactivation and anti-proliferation of T cells and B cells, selective increase of functional regulatory T cells, and inhibition of antibody production [20]. Importantly, it inhibits fibroblasts proliferation [21]. Sirolimus has been used in clinical practices for coating coronary stents [22], preventing rejection in organ transplantation [23], and treating autoimmune diseases [24-26], such as lymphoproliferative syndrome, systemic lupus erythematosus (SLE) and primary antiphospholipid syndrome. However, sirolimus has not been reported to treat RPF.

For investigating if sirolimus would be a potential treatment option for idiopathic RPF, we searched for the gene targets of sirolimus in three drug databases, SwissTargetPrediction [27], DrugBank [28], and CHEMBL [29]. Collectively, over 400 affected genes were identified, and even though the overlap among the three databases was little, the enriched GO terms and KEGG terms were common (Supplementary Figure 2). For systematically evaluating the overlap between the biological terms affected by sirolimus and RPF, i.e., pharmacology vs pathology, we developed a paring score, in which the top 50 terms from each GO enrichment set (i.e., pharmacological terms vs pathological terms) were compared. This comparison took a tiering strategy, in which the top 20 terms were assigned as tier 1, and the 21st-50th terms were assigned tier 2. Matching between tier 1 terms from both data sets would be assigned a higher weight; whereas matching between tier 1 terms from one dataset and tier 2 terms from the other dataset would be assigned a reduced weight. Summing up the weights results in the paring score (Method). A large overlap was observed for the sirolimus pharmacology and RPF pathology (Supplementary Figure 3A), with a pairing score of 0.70 (Supplementary Table 1). We also constructed the overlap for prednisone and tamoxifen, the current and the previous generations of treatment for RPF, respectively. Prednisone had relatively fewer overlaps with a pairing score of 0.58 (Supplementary Figure 3B). Not surprisingly, tamoxifen had the least overlap among the three drugs, with a pairing score of 0.43 (Supplementary Figure 3C). Therefore, we concluded from this pathway analysis that sirolimus had a high potential for treating RPF.

**Figure 2.**
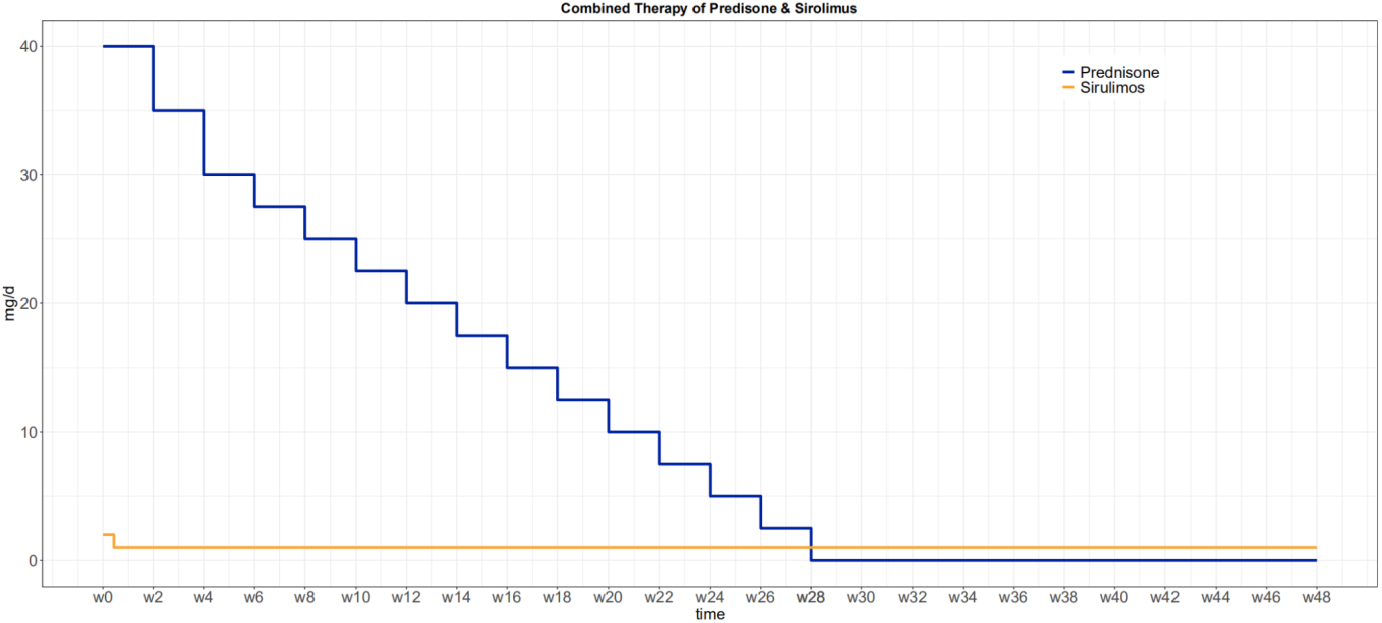
A prescription of prednisone and sirolimus as a combined therapy for treating RPF in an individual of 50 kg of weight.

**Figure 3.**
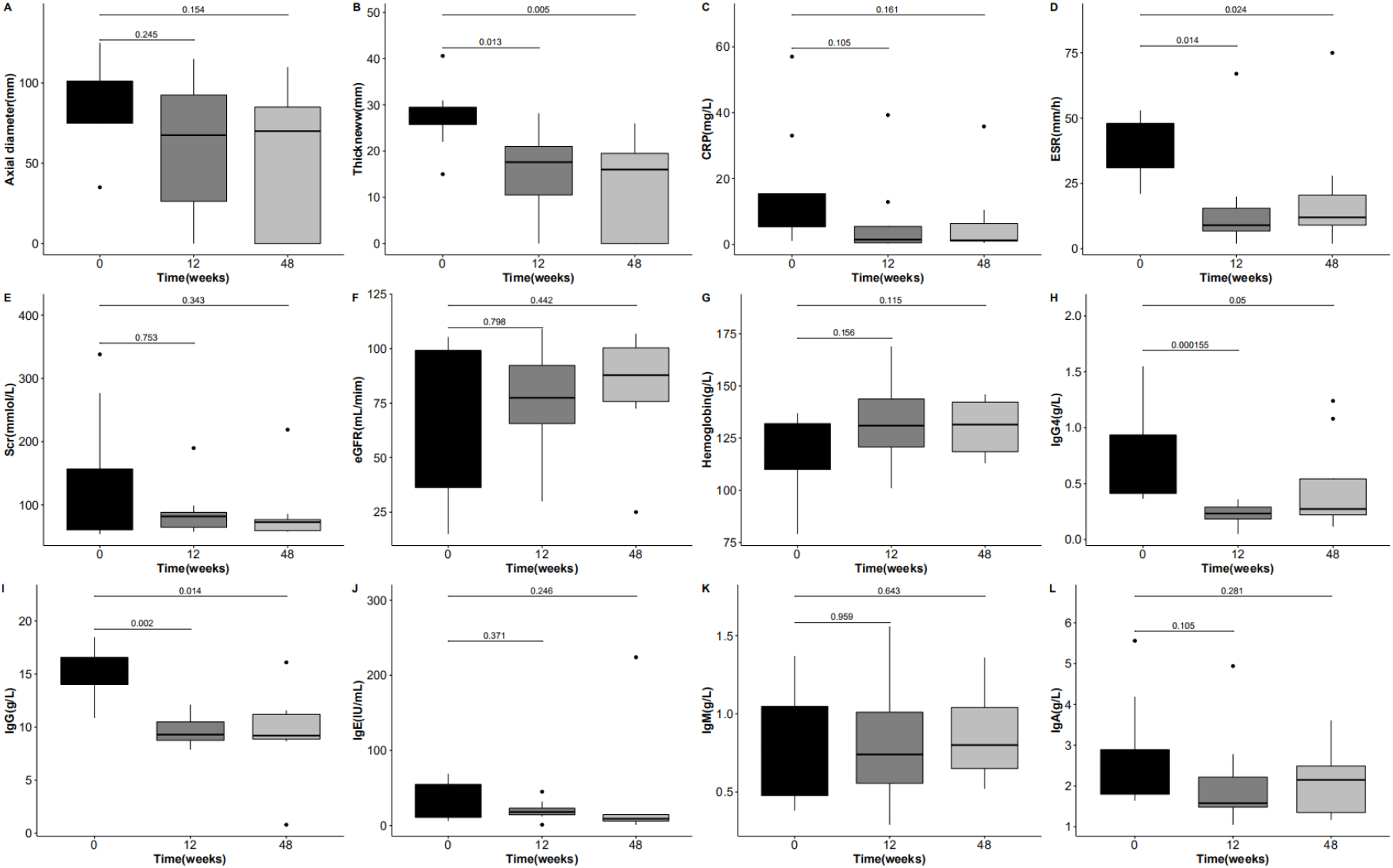
Lab tests to assess the treatment outcomes of prednisone and sirolimus combined therapy for RPF. Displayed are **(AB)** Changes of fibrous tissue mass, **(CD)** Inflammatory markers, **(EF)** kidney functions, **(G)** hemoglobin, **(H)** IgG4, and **(I-L)** Immunoglobulin major types, as measured in all 8 patients with the combined therapy.

### Design of a combined therapy with prednisone and sirolimus to treat RPF

We next implemented a clinical trial to evaluate the actual effects of combining prednisone and sirolimus in treating RPF. Referencing the current clinical practices of prednisone mono-therapy, we designed a two-phase treatment strategy:

① Prednisone acetate at 0.8 mg/kg/day (maximum dosage 60 mg/day), reduced by 5 mg every 14 days until reaching 30 mg/day, and reduced by 2.5 mg every 2 weeks until discontinuation.

② Sirolimus at 2 mg/day for the first three days and around 1 mg/day thereafter, with plasma drug concentration monitored at 2 weeks, 12 weeks, and 48 weeks of treatment to maintain a stable level at 4-15 ug/L.

An illustrative prescription protocol is given in Figure 2 for an individual of body weight of 50 kg. In phase I, both prednisone and sirolimus were prescribed as aforementioned. In phase II, sirolimus was used as a monotherapy for maintaining the treatment effects. Assessments were collected at the baseline (i.e., onset of the treatment), 12 weeks and 48 weeks of treatment, including contrast-enhanced computed tomography (CT) to determine sizes of the fibrous tissues, lab tests on inflammation, kidney and liver functions, documentation of side effects, cell types and abundance by flow cytometry, and circulating inflammatory proteins in plasma by proteomics.

### Patient characteristics

In total we recruited twelve RPF patients for the combined therapy trial. Eight patients, including five females and three males, completed the 48-week long assessments. The rest four people dropped out due to the reason for not following the prescriptions strictly. Patient characteristics at baseline were documented in Supplementary Table 2. In each patient, one fibrous tissue locus was observed. The median thickness of the fibrous mass was 28.85 mm, and the craniocaudal length was 92.50 mm (Supplementary Table 3). Five patients developed hydronephrosis and four of them had ureteral stents implanted to alleviate the symptom (Table 1). On average, the combined intake of prednisone and sirolimus lasted for 30.8 ± 3.5 weeks, followed by the sirolimus monotherapy for 17.2 ± 3.5 weeks.

### RPF symptoms were improved with the combined therapy

Along the treatment, most of the RPF symptoms displayed improvement (Supplementary Table 3, Figure 3). CT scan showed that the fibrous tissue shrunk nearly by half in axial diameter and thickness at both 12 weeks and 48 weeks of treatment. Two markers of acute inflammation, C-reactive protein (CRP) and erythrocyte sedimentation rate (ESR), were significantly reduced. Two markers of kidney functions, serum creatinine (Scr) and the estimated glomerular filtration rate (eGFR), displayed constant improvement along the treatment, with the values after treatment aligned to the normal range. The combined therapy also corrected for anemia in RPF patients, with hemoglobin concentrations raised by about 14%. Interestly, the circulating levels of IgG4 displayed a significant reduction along the treatment. We also profiled the major isotypes of immunoglobulins, and found that both IgG and IgE antibodies were reduced along the treatment while IgM and IgA did not show any significant changes.

### Profiles of immune cell types and abundances

We performed comprehensive immunophenotyping by flow cytometry on 7 subsets of T cells in peripheral blood (Methods, Supplementary Figure 4). Over the course of the therapy, a trend of reduction in cell abundance was observed for T_H_2, T_H_17 and circulating T follicular helper cells-like cells (cT_FH_-like cells) (Figure 4). T_H_2 and T_H_17 cells are IL-4-producing and IL-17-producing CD4+ T cells, respectively. cT_FH_-like cells (T follicular helper cells) are known to play important roles in the development of GCs as well as antibody production. Their upregulation has been reported in other autoimmune conditions such as SLE [30, 31].

**Figure 4.**
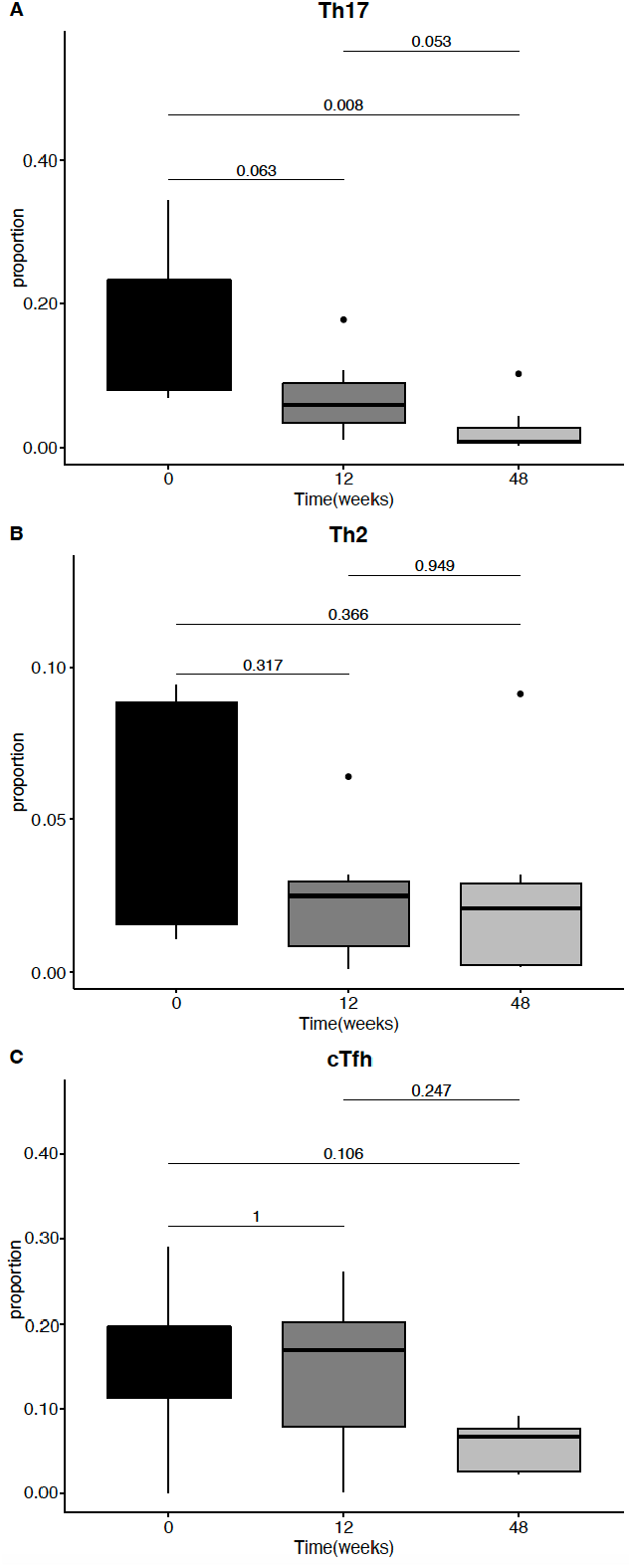
Immunoprofiling by flow cytometry indicated that specific T cell subsets were regulated with the therapy. Changes were in proportions for **(A)** Th2 cells, a type of IL-4 producing CD4+ T cells, with the markers of CD3^+^CD4^+^CD8^−^CXCR3^+^CCR6^−^CCR4^+^CCR7^low^ ; **(B)** Th17 cells, a type of IL-17 producing CD4+ T cells, with the markers of CD3^+^CD4^+^CD8^−^CXCR3^−^CCR6^+^CCR4^+^CCR7^lo^; **(C)** circulating T follicular helper cells-like cells (cT_FH_ -like cells), with the markers of CD3^+^CD4^+^CD8^−^CD45RA^−^CXCR5^+^CCR7 ^low^PD-1^high^.

### Profiles of circulating cytokines

We leveraged the Olink protein biomarker platform to systematically quantify circulating cytokines in the RPF patients with the combined treatment versus in the age- and sex-matched healthy controls. In particular, oligonucleotides coupled with antibodies to the inflammatory cytokines were amplified and sequenced using the next-generation sequencing technology, and therefore low abundant cytokines, even at the level of sub-picogram per milliliter, could be accurately measured [32]. Out of the 96 targeted inflammatory cytokines, we obtained qualification for 76 of them (Methods). A total of 28 cytokines displayed significant changes in diseased state relative to the healthy controls, hence “abnormal cytokines” (t-test, FDR corrected P < 0.05, Supplementary Figure 5). These included: tumor necrosis factor (TNF) and related proteins, such as TNFRSF9, TNFSF14, TRAIL (TNF-related apoptosis-inducing ligand), and CD40; chemokines such as CCL3, CCL19 and CCL23; interleukin (IL) cytokines and related proteins, such as IL-6, IL-8, IL-12B, IL-18, and IL-10RB, and other proteins. After treatment (Figure 5A), 5 of these abnormal cytokines regressed towards healthy levels, namely TNF, TNFRSF9, TNFSF14, CCL23 and IL-12B (t-test, FDR corrected P < 0.05). Furthermore, the therapy caused 9 additional cytokines to fluctuate significantly, hence “treatment responsive normal cytokines”. Although they were termed “normal” due to no significant change in diseased state, a careful examination revealed that 5 such cytokines, namely TWEAK (TNF-like weak inducer of apoptosis), SCF (stem cell factor), Flt3L (Fms-like tyrosine kinase 3 ligand), CCL11 and CCL28, displayed marginally reduced levels in diseased state, although they did not pass the threshold of statistical significance; the treatment brought them closer to the healthy levels. For the other 4 cytokines, CCL25, IL-17C, ST1A1 and LIF-R, no trend of difference between cases and controls were observed, therefore these perturbations were likely solely attributed to drug side effects. We performed KEGG pathway enrichment on the 14 treatment responsive cytokines, and found that the most altered pathways were cytokine and chemokine signaling pathways (Figure 5B).

**Figure 5.**
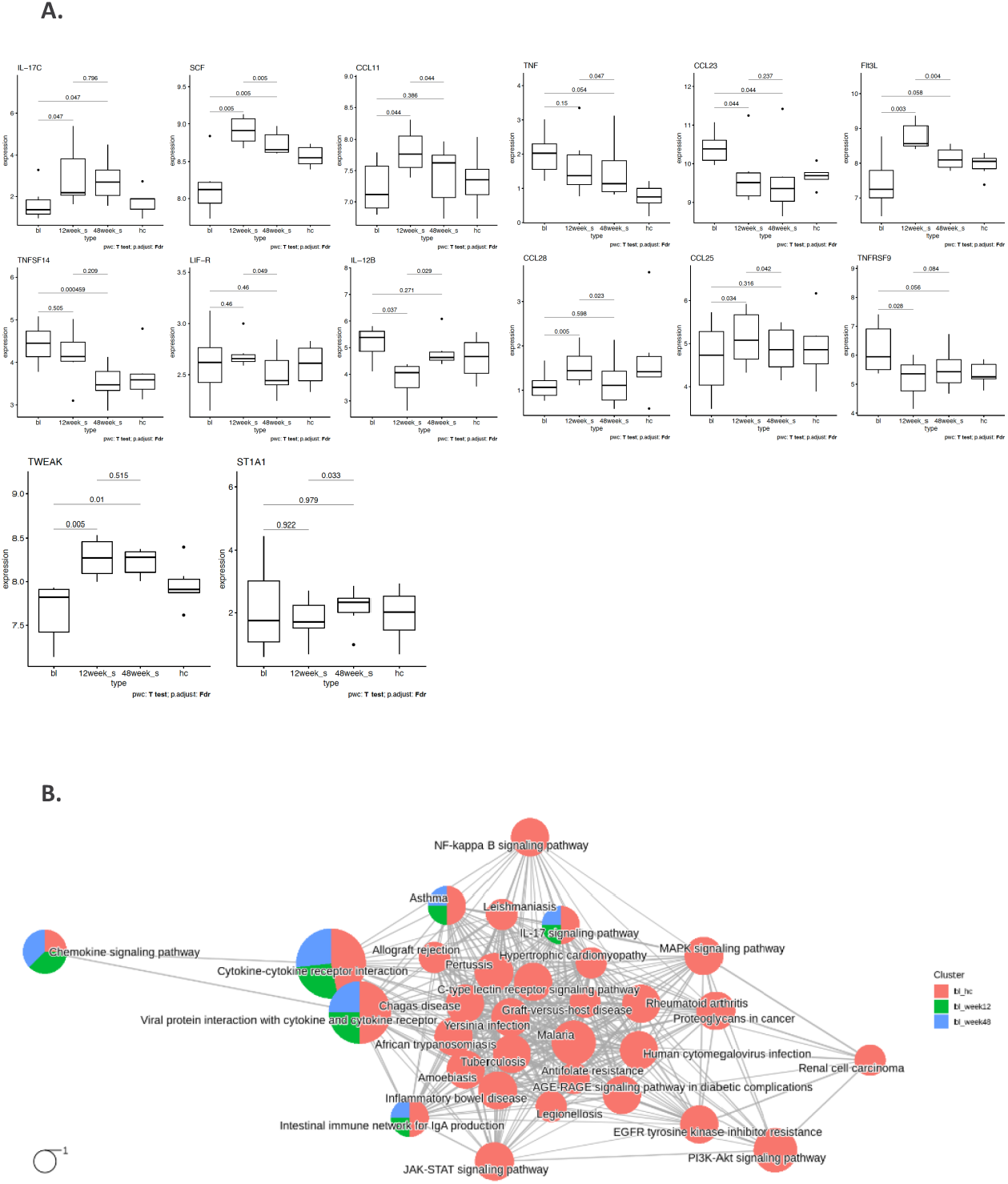
Cytokine profiling via Olink Target 96 inflammation panel. **(A)** Cytokines and chemokines with significant changes after treatment were displayed. In total 14 proteins were identified. **(B)** Signaling pathways enriched in the Olink proteins that were shown with significant changes in various conditions. Red: changed in baseline versus healthy controls; green: changed in baseline versus after 12-week treatment; blue: changed in baseline versus after 48-week treatment.

### Side effects of the combined therapy

We recorded a few adverse reactions during phase I of the therapy, when both prednisone and sirolimus were prescribed: 6 patients with hyperlipidemia, 2 patients with hyperglycemia, 1 patient with hypertension and 1 patient with muscle soreness (Table 1). After prednisone was removed in phase II, most of these adverse effects were gone, except for a female patient who developed Herpes Zoster, and a male patient whose hyperlipidemia and muscle soreness remained. Overall, the combined therapy was tolerant, and it was evident that side effects of the steroid hormone were much reduced in Phase II of the therapy, when prednisone was removed and sirolimus became the single drug to maintain the treatment effects.

## Discussion

In this study, we presented a combined therapy of prednisone and sirolimus to treat idiopathic RPF as opposed to the classic steroid hormone therapy. It leveraged the discovery that mTOR was highly activated in RPF tissues, and the biological pathways affected by sirolimus had a high degree of match with that in RPF (paring score of 0.70). Our strategy was to use both prednisone and sirolimus in phase I, in which a regular dosage of prednisone was used for a boost start to initiate fibrosis reduction, followed by a gradually reduction while maintaining a stable plasma concentration of sirolimus; when prednisone was reduced to none, the treatment entered phase II, in which sirolimus remained the only drug to sustain the fibrous remission. In our clinical trial in RPF patients over a one-year period, we observed that this strategy shrunk fibrous tissues, reduced inflammation, and improved kidney function and anemia. These improvements were evident in phase I and remained at similar levels in phase II, suggesting sirolimus was able to maintain the treatment effects. It is interesting to note that although most RPF patients in our cohort had relatively low levels of IgG4 (i.e. only one of the eight patients was classified as IgG4-RD), the combined therapy did further bring down their IgG4 levels, suggesting it was also potent for treating the IgG4^+^ RPF. Furthermore, most side effects induced in phase I, such as hyperlipidemia and hyperglycemia, disappeared in phase II, suggesting side effects of the steroid hormone were temporary and controlled in this therapy. Overall, our new treatment strategy of using gradient reduction of prednisone and a long-term stable dosage of sirolimus demonstrated by our single-arm one-year clinical trial to be effective for treating idiopathic RPF with tolerant side effects. As the current treatments of RPF involve long-term usage of steroid hormones and often resulted in severe side effects, our therapy seems to alleviate this issue, casting hope for a new treatment direction.

Treatments to idiopathic RPF rely on medication to suppress inflammation and relief of ureteral obstruction via e.g. stent implantation, percutaneous nephrostomy and ureterolysis. For medication, the first generation drug was tamoxifen, which has long been used as an anti-estrogen compound for treating early-stage, estrogen-sensitive breast cancer and for treating immune disorders in the recent three decades [33]. Tamoxifen alters immune profiles by inducing a shift from cellular (T-helper 1) to humoral (T-helper 2) immunity, and demonstrated to exert anti-inflammatory and anti-fibroblastic functions in RPF, whose long-term safety was proven by van Bommel et al [34]. Later in 2011, Vaglio *et al* reported in a randomized clinical trial that glucocorticoid monotherapy achieved better treatment outcomes over tamoxifen [9], and has since become the mainstay therapy for RPF. Indeed, glucocorticoids induced rapid fibrosis remission and retreat of acute-phase inflammation. Using computational approaches, we quantified the matching between the drug effects (as recorded in drug-gene databases) and the RPF pathological pathways (as recorded in the gene-disease databases), and found that the paring scores were 0.43 for tamoxifen-RPF, 0.58 for prednisone-RPF, and 0.70 for sirolimus-RPF, displaying a trend of continuous improvement. This indicated that sirolimus, as an mTOR inhibitor, more specifically captured the altered biological pathways in RPF.

Our study suggested mTOR as a critical player in RPF. Indeed, mTOR emerged as an important common signaling molecule in fibrosis, which promotes fibroblast proliferation and strengthens pro-inflammatory responses via proliferating Th1, Th17 and CD4^−^CD8^−^ T cells. mTOR inhibition by sirolimus was demonstrated to improve numerous autoimmune diseases, including SLE, Refractory Lupus, juvenile idiopathic arthritis and Sjogren’s Syndrome [35-37]. Our clinical trial suggested sirolimus combined with prednisone as a promising therapy for treating RPF, expanding the list of autoimmune conditions that could benefit from mTOR inhibition. However, it is worth noting that not all autoimmune conditions involving fibrosis are treatable by sirolimus. A previous follow-up study on SSc and sirolimus treatment suggested a limited treatment effect, although mTOR activation was evident in SSc, leading to production of type I collagen in fibroblasts, a critical step of fibrosis formation [38]. Clinical trials and careful examinations are needed to evaluate the drug effects.

Fibrosis is a common complication of diseases and accounts for up to 45% of death in industrialized countries [39]. Although no effective medicine has existed yet to completely revert the fibrous process, it is now known as highly dynamic and therefore presents opportunities for correction and cure. The severe form of RPF results in uterus obstruction, kidney failure, and encasement of abdominal aorta and iliac aorta. Containing the fibrous mass development and reducing the existing fibrosis are crucial, which usually involves long-term uptake of medicine. A safe therapy with minimal side effects is therefore strongly desired. As mTOR signaling is general to fibrosis, the strategy of combining a much reduced dosage of steroid hormones and a stable dosage of sirolimus may have a broad application to other fibrosis-related diseases.

There are several limitations in the current study. First of all, the patient cohort size was small, limited to eight patients. As RPF is a rare condition with a low prevalence rate, i.e. about 1.4 cases/100,000 inhabitants [2], patient recruitment has been challenging. Secondarily, randomized clinical trials are desired to evaluate treatment differences between our proposed combined therapy and the standard prednisone monotherapy. For this we are carrying out a randomized clinical trial (NCT04047576) and in the process of collecting data for analysis. A rigorous examination will provide quantified and more solid information of the benefits of using the 2-phase sirolimus-prednisone combined therapy versus the prednisone monotherapy. Thirdly, our plasma proteomic assays indicated that half of the abnormal cytokines and chemokines (14 out of the 28 perturbed proteins) displayed a trend towards healthy levels after the treatment, and a small number of proteins at normal levels displayed perturbation after the treatment (4 out of the 76 quantified proteins, about 5%). As only 76 inflammation-related proteins were quantified and some of the key players including IL-4, IL-13 and IL-21 were not detected, our detection power was limited. A more comprehensive treatment assessment will be revealed by including a larger number of proteins. Future studies with more RPF patients, longer follow-up periods and more inclusive measurements will provide richer information and stronger analytical power for assessing new treatment strategies, including our proposed therapy of combining prednisone and sirolimus.

## Methods

### Patient Recruitment

Idiopathic RPF patients were enrolled from Peking University International Hospital. Inclusion criteria were: 1) 18-75 years old; 2) diagnosed as idiopathic RPF by CT or magnetic resonance imaging (MRI) - for patients suspected to have secondary RPF or atypical idiopathic RPF, puncture biopsy was conducted for clarification; and 3) increased ESR and CRP levels caused by RPF or active lesions detected by imaging. Exclusion criteria were: 1) secondary retroperitoneal fibrosis; 2) usage of any glucocorticoid (equivalent to >10 mg per day of prednisone), immunosuppressant, or biologic medication within 3 months prior to the enrollment; 3) having any contraindication of glucocorticoid or sirolimus, allergic to sirolimus, or experienced serious adverse reactions from previous use of any of the above drugs; 4) massive proteinuria (24-hour urine protein quantitation ≥3 g), moderate-to-severe anemia (hemoglobin <90 g/L), agranulocytosis (white blood cell count <1.5×10^9^/L or neutrophil count <0.5×10^9^/L), platelet count < 50,000, or interstitial pneumonia; 5) uncontrollable diabetes, hypertension, hyperlipidemia, infection, or heart failure; 6) having any malignant tumor; 7) other serious complications or general conditions that do not permit study enrollment; 8) pregnancy or plan for pregnancy in the near future; or 9) unable to adhere to follow-up or refuses to provide consent.

The study was performed in accordance with the Declaration of Helsinki. The study was approved by the Research Ethics Committee at Peking University International Hospital (ethics approval number: 2019-031 [BMR]). All patients provided written informed consent.

### Clinical study design

The combined therapy evaluated in this study consisted of two phases. In phase I, a gradually decreasing dosage of prednisone acetate was prescribed to the patients together with sirolimus. In phase II, prednisone was discontinued and only sirolimus was prescribed. The dosages were arranged as follows: prednisone was given at 0.8 mg/kg/day (maximum dosage 60 mg/day), reduced by 5 mg every 14 days until reaching 30 mg/day, and further reduced by 2.5 mg every 2 weeks until reaching zero. Sirolimus was given at 2 mg/day for the first three days and 1 mg/day thereafter, with plasma drug concentration monitored at 2 weeks, 12 weeks, and 48 weeks of treatment to maintain a stable level at 4-15 ug/L.

Treatment failure and relapse were assessed according to a randomized controlled trial for RPF [9]. Treatment failure was defined as meeting at least two of the three criteria: 1) symptoms were not relieved; 2) ESR and CRP levels remained abnormal and have fallen to less than 30% compared to the baseline levels; and 3) the retroperitoneal fibrous mass did not shrink, i.e., shrinkage of the maximal cross-sectional area was < 50% or the size even increased. If symptoms caused by infection or other factors did not relieve or any inflammatory marker was abnormal, re-examination and evaluation were conducted 4 weeks after the inducing factors were controlled. Relapse was defined as meeting at least one of the following three criteria during the follow-up period: 1) recurrence or new-onset disease-related symptoms, 2) hydronephrosis, or 3) fibrous tissue enlarged by 20% (maximum cross-sectional area) as measured by CT or MRI. For patients who displayed only disease-related symptoms but did not have hydronephrosis or fibrous mass enlargement, relapse was confirmed with a 50% increase in ESR or CRP values compared to that recorded at the previous visit [9].

### Laboratory tests

At the time of study enrollment (baseline, i.e. without treatment), 12 weeks and 48 weeks of treatment, the patients underwent physical examination, abdominal CT and peripheral blood sample collection. Laboratory tests were performed on the peripheral blood, including complete blood count (CBC), erythrocyte sedimentation rate (ESR), C reactive protein (CRP), serum immunoglobulin (Ig) level, Immunoglobulin G4 (IgG4) level, serum creatinine and estimated glomerular filtration rate (e-GFR), liver function tests, serum electrolytes, fasting glucose, lipid profile, and urinalysis.

Side effects were monitored every 1-3 months as appropriate by a checklist of standardized items, blood pressures and the aforementioned laboratory tests.

### Immunohistochemical assays

RPF tissue samples were derived from 16 patients with active RPF, with the fibrous mass derived by puncture biopsy or surgery. Control samples were derived from the mesenteric root tissue from 4 early-diagnosed colon cancer patients who were confirmed by pathological examination to have no lymphatic metastasis at the time of biopsy. Tissue samples were fixed in 10% neutral buffered formalin, embedded in paraffin and cut into slices of 4 μm thick. All tissue sections were routinely stained with Hematoxylin-Eosin and incubated at 58°C for 1 h, followed by deparaffinization and rehydration with xylol and graded ethanol. After antigen retrieval, non-specific antigen sites were blocked and tissue sections were incubated with mTOR antibodies (Abcam, dilution 1:400), P-AKT (S473) antibodies (Abcam, dilution 1:500) and P-S6K1 (T389+T412) antibodies (Abcam, dilution 1:100). The primary antibodies were revealed with the appropriate secondary antibodies (Abcam). Peroxidase activity was revealed by 3-30-diamino-benzidine-tetrahydrochloride (DAB). The fibrotic area was manually outlined, and the software Visiopharm Integrator System was used for quantification.

### Immune profiling by flow cytometry

All 8 RPF patients consented to donate blood for comprehensive immunological analysis. Protocol specific immunophenotypic analysis of peripheral blood leukocyte subsets of patients was performed at baseline, week 12 and week 48. PBMCs were isolated from blood samples by the Percoll gradient density centrifugation. Cell staining was performed with fluorescence conjugated antibodies against cell surface markers at room temperature for 30-40 minutes. All antibodies for flow cytometry in this study are listed in Supplementary Table 4. Proportions of regulatory T cells (Treg), Mucosal-associated invariant T cells (MAIT), circulating T follicular helper-like cells (cTfh), T follicular regulatory cells (Tfr) and T helper 1 cells (Th1), T helper 2 cells (Th2), and T helper 17 cells (Th17) were acquired on a Beckman Coulter Cytoflex LX flow cytometer and analyzed by FlowJo (version 10.8.1). The cell type markers were listed in Supplementary Figure 4B.

### Olink proteomics assays

Intravenous blood samples were collected. EDTA anticoagulation was performed, followed by centrifugation at 3000 rpm for 15 min. Supernatant containing proteins was collected for Olink proteomic analysis, following standard protocol by Olink. Briefly, Cytokines and chemokines were profiled using the proximity extension assays (PEA) in a 96-plex inflammatory panel developed by Olink Proteomics (Sweden) and serviced by the Shanghai Biotechnology Corporation. Standard protocols for quality control and data normalization by referencing internal and external controls were carried out in the Olink NPX Manager software (version 3.3.2.434). The normalized protein expression (NPX) values, as a relative quantification method, was used for comparing expression levels of individual proteins in different conditions.

### Bioinformatics

For pathologic analysis, genes affected in RPF were obtained by querying the key word “retroperitoneal fibrosis” in the databases including MalaCards, DisGenet and GeneCards. For pharmacologic analysis, genes affected by the drugs sirolimus, prednisone and tamoxifen, respectively, were obtained by querying the databases including SwissTargetPrediction, DrugBank, and CHEMBL. All gene set enrichment analysis, including those from Olink proteomics, was performed via ClusterProfiler (version: 4.4.4) and adjusted P value threshold was set at 0.05.

For comparing the enriched terms between the pathologic and pharmacologic gene sets, a pairing score was developed, in which the top 20 enriched terms from one gene set were matched with the top 50 enriched terms from the other gene set. The matching was classified into two tiers. Tier 1 refers to matching the top 20 terms in the other gene set, with each match given full weight of 1. Tier 2 refers to matching the top 30-50 terms in the other gene set, with each match granted the weight of 0.5. The overall paring score was the sum of the tier 1 and tier 2 scores, generalized as:

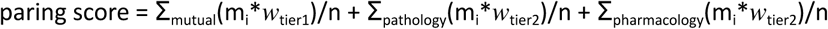

where, m_i_ denotes a matching event between pathology terms and pharmacology terms; n is the number of terms classified as tier 1, set as 20 in our analysis; *w*_tier1_ is the weight for any matching in tier 1 terms, set as 1; *w*_tier2_ is the weight for any matching in tier 2 terms, set as 0.5.

### Statistical Analysis

Lab test results and flow cytometry quantification results were assessed by wilcoxon rank-sum tests. Cytokine and chemokine expression levels measured by Olink were assessed by t-test, in which the RPF case group to healthy control group comparison was performed by group-level t-test, and the RPF treatments versus baselines were compared by paired t-test. Instead of using a nominal p-value of 0.05, FDR corrected p-value was used as the new threshold.

## Data Availability

All data produced in the present study are available upon reasonable request to the authors

## Acknowledgment

We thank Dr. Shengguang Li from Peking University International Hospital, Dr. Jinxia Zhao from Peking University Third Hospital, Dr. Xiaoying Zhang from Peking University People’s Hospital for referring study participants; Dr. Feng Yu from the Nephrology Department of Peking University International Hospital for assistance in immunohistological experiments on tissue biopsies; Dr. Tong Zhang from the Pathology Department from Peking University International Hospital for assistance in pathological analysis; Drs. Jing Li and Xiaoying Zhang from Peking University International Hospital for collecting peripheral blood samples. We also thank members of the Laboratory of Intelligent Computing in Biomedicine in the Greater Bay Area Institute of Precision Medicine (Guangzhou) for insightful discussions and suggestions.

