## Supplementary Figures and Tables for "Combined therapy of prednisone and mTOR inhibitor sirolimus for treating retroperitoneal fibrosis"

**Affiliations:**
1. Department of Rheumatology and Immunology, Peking University International Hospital, Beijing, China
2. Department of retroperitoneal tumor surgery, Peking University International Hospital, Beijing, China
3. Center for Intelligent Medicine Research, Greater Bay Area Institute of Precision Medicine (Guangzhou), School of Life Sciences, Fudan University. Guangzhou, China
4. Department of Pharmacy, Peking University International Hospital, Beijing, China
5. Department of Rheumatology and Immunology, Peking University People's Hospital, Beijing, China

* These authors contributed equally to this work.

**# Correspondence:**
Chenghua Luo:
Cuiping Pan:

### Supplementary Figures

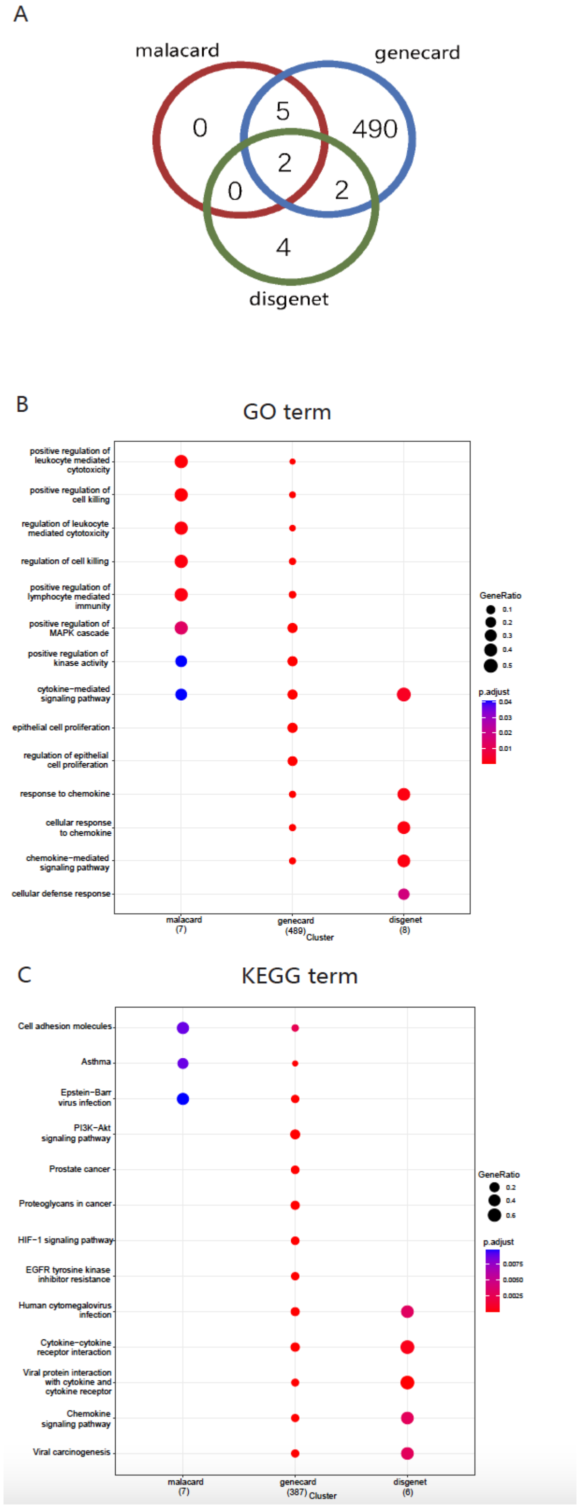


**Supplemental Figure 1**. Pathology pathway enrichments for RPF. **(A)** Overlap of genes queried from the three gene-disease databases; **(B)** Gene Ontology (GO) enrichment for the identified genes in each database; **(C)** Pathway enrichment using Kyoto Encyclopedia of Genes and Genomes (KEGG) database for the identified genes in each database.

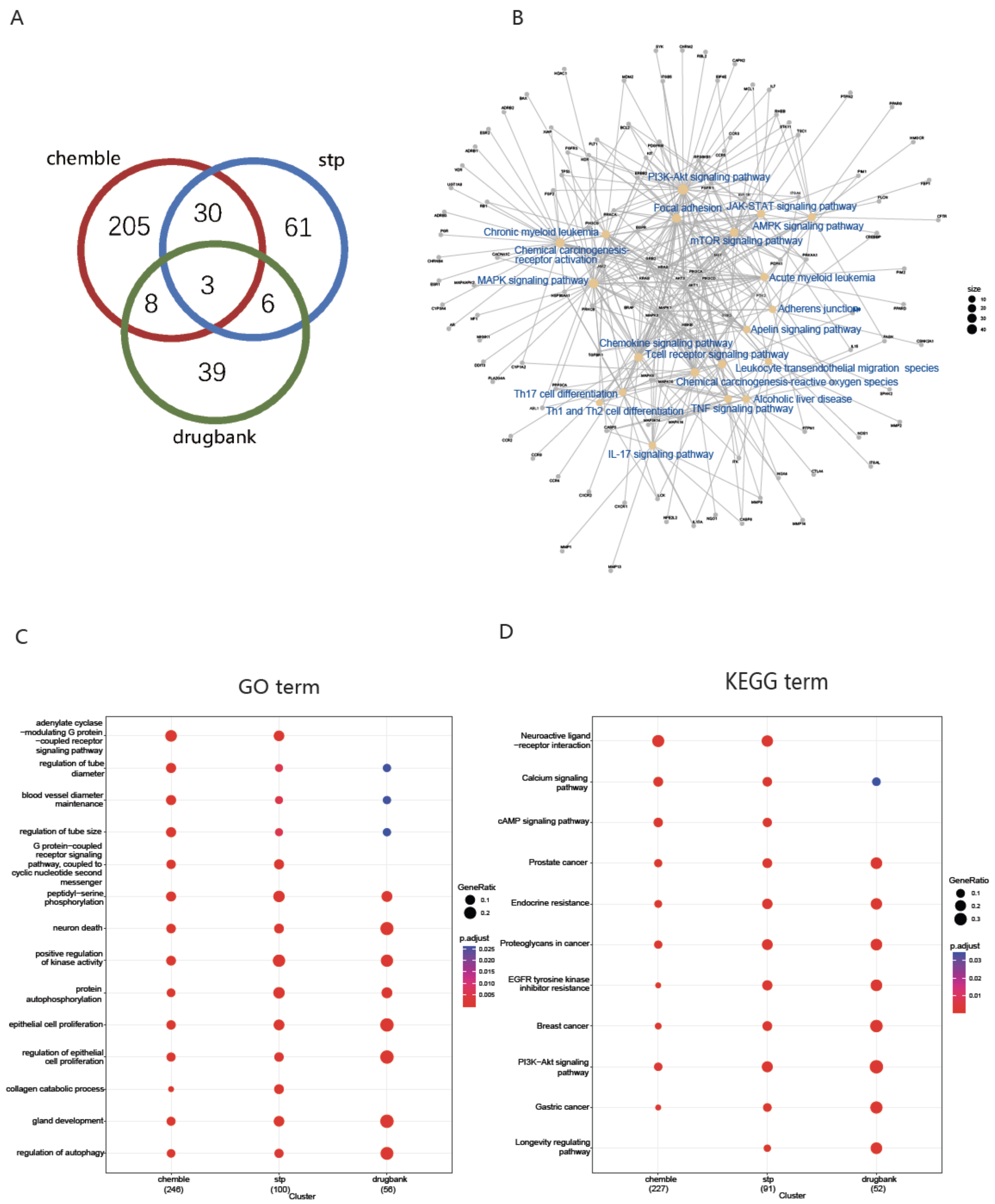


**Supplemental Figure 2.** Target genes and pathways of sirolimus. **(A)** Overlap of genes queried by sirolimus from the three drug-gene databases; **(B)** signaling pathway network enriched from genes affected by sirolimus. **(C)** GO enrichment for the identified genes in each database; **(D)** Pathway enrichment using KEGG database for the identified genes in each database.

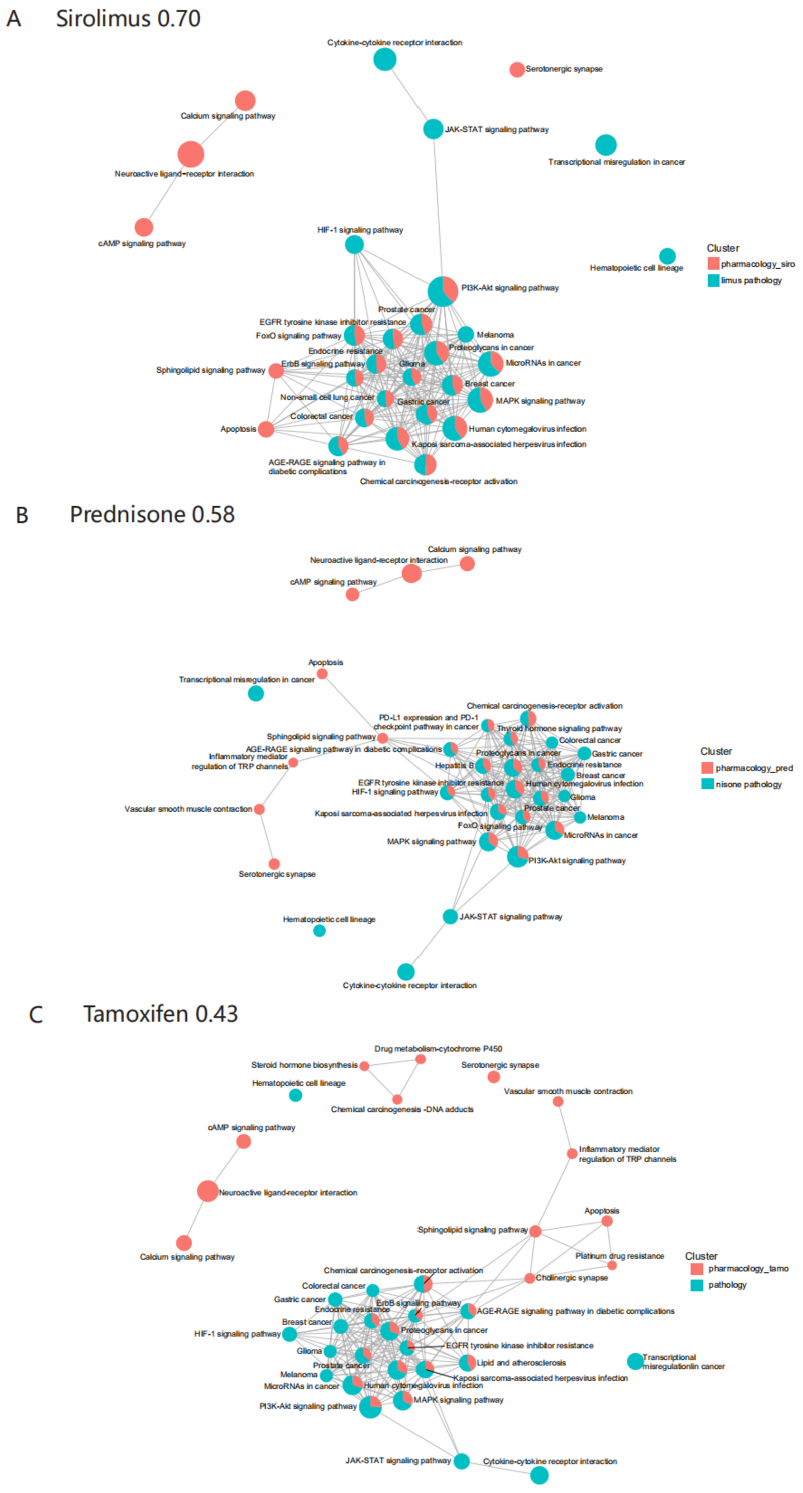


**Supplemental Figure 3. Biological pathways affected by the drugs and by RPF, with those for** sirolimus in **(A)**, prednisone in **(B)** and Tamoxifen in **(C)**. Pathways affected by the drugs are indicated in red, and the pathways affected in RPF are indicated in cyan. The proportion in the pie chart is based on the number of detected genes in that particular pathway. Pairing scores indicating the matchness between the drug and the RPF pathology are listed next to the drug name.

A

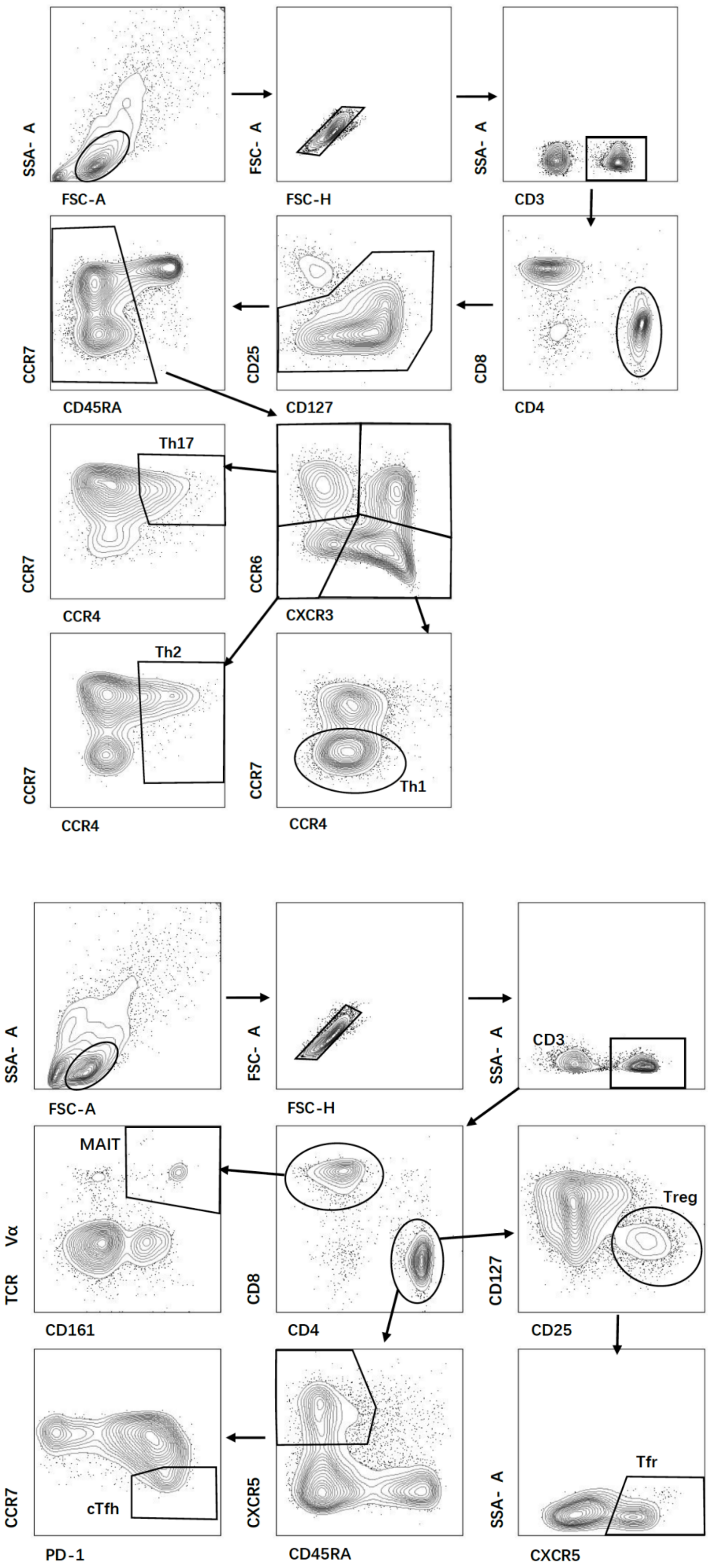

SSA- A
FSC-A
FSC- A
FSC-H
SSA- A
CD3
CCR7
CD45RA
CD25
CD127
CD8
CD4
Th17
CCR7
CCR4
CCR6
CXCR3
Th2
CCR7
CCR4
CCR7
CCR4
Th1
SSA- A
FSC-A
FSC- A
FSC-H
SSA- A
CD3
MAIT
TCR Vα
CD161
CD8
CD4
Treg
CD127
CD25
cTfh
CCR7
PD-1
CXCR5
CD45RA
Tfr
SSA- A
CXCR5

B

| | |
|---|---|
| Th1 | $CD3^+CD4^+ CD8^-CXCR3^+CCR6^-CCR4^-CCR7^{low}$ |
| Th2 | $CD3^+CD4^+CD8^-CXCR3^+CCR6^-CCR4^+CCR7^{low}$ |
| Th17 | $CD3^+CD4^+CD8^-CXCR3^-CCR6^+CCR4^+CCR7^{low}$ |
| Treg | $CD3^+CD4^+ CD8^-CD25^{high}CD127^{low}$ |
| MAIT | $CD3^+CD4^-CD8^+TCRV\alpha^+CD161^{high}$ |
| cTfh | $CD3^+CD4^+CD8^-CD45RA^-CXCR5^+CCR7^{low}PD\text{-}1^{high}$ |
| Tfr | $CD3^+CD4^+CD8^-CD25^{high}CD127^{low}CXCR5^+$ |

**Supplementary Figure 4. Phenotypic characterization of T-cell subsets by Flow cytometry. (A)** Illustrative examples of the lymphocyte gating strategy for identifying T cell subsets; **(B)** A complete marker list for each T cell subset.

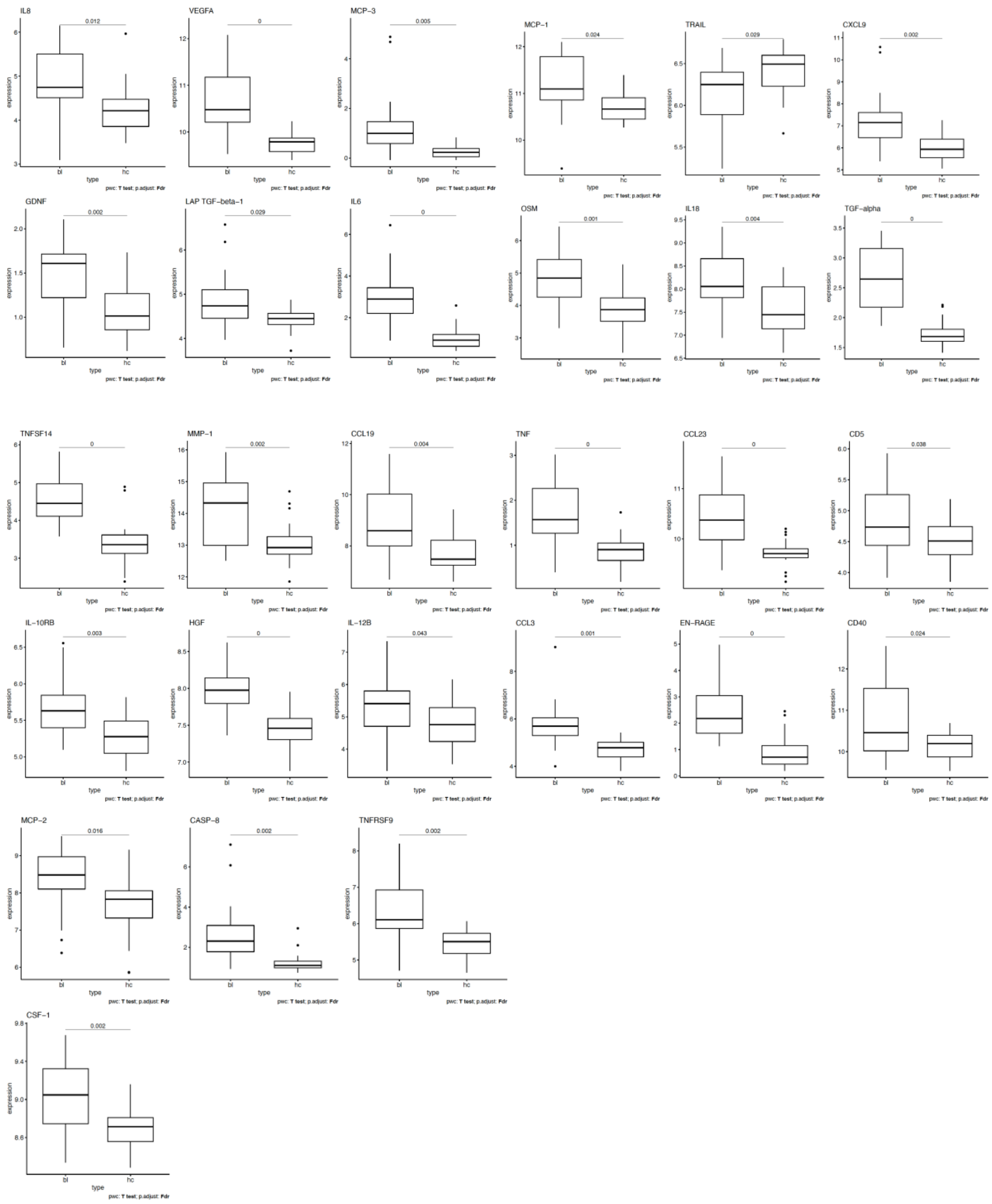


**Supplementary Figure 5. Cytokines displaying significant changes between RPF patients (bl) and age- and sex- matched controls (hc).** Bl: baseline; Hc: healthy controls.

### Supplementary Tables

**Supplementary Table 1. Pairing scores for evaluating matching between disease pathology and drug pharmacology.**

| Overlap of Enrichment Terms | | | | | | |
|---|---|---|---|---|---|---|
| Catertory | pharmacology_sirolumus | pathology | pharmacology_prednisone | pathology | pharmacology_tamoxifen | pathology |
| Input | 50 | 50 | 50 | 50 | 50 | 50 |
| Tier-1 overlap | 10 | 10 | 7 | 7 | 4 | 4 |
| Tier-2 overlap | 4 | 4 | 5 | 4 | 3 | 6 |

| Paring Scores | | | |
|---|---|---|---|
| Catertory | sirolimus vs RPF | prednisone vs RPF | tamoxifen vs RPF |
| Tire-1 | 0.50 | 0.35 | 0.20 |
| Tier-2 pharmacology | 0.10 | 0.13 | 0.08 |
| Tier-2 pathology | 0.10 | 0.10 | 0.15 |
| Total | 0.70 | 0.58 | 0.43 |

**Supplementary Table 2. Baseline characteristics of the patients.** Lab measurements are presented in median values, followed by 25th-percentile and 75th-percentile values in parenthesis. The category of "Pathological features" refers to $IgG4^+$ status.

| Clinical Features | Counts or lab measurements | Percentage |
|---|---|---|
| **Sex** | | |
| Male | 3 | 37.5% |
| Female | 5 | 62.5% |
| **Age** | 57.0 (52.0, 64.5) | |
| **Time before treatment (months)** | 2.5 (1.1, 21.0) | |
| **Symptoms** | | |
| Back pain | 5 | 62.5% |
| Abdominal pain | 2 | 25.0% |
| Lower limb edema | 1 | 12.5% |
| Constipation | 1 | 12.5% |
| **Hydronephrosis** | | |
| Overall | 5 | 62.5% |
| Left | 1 | 20.0% |
| Right | 1 | 20.0% |
| Bilateral | 3 | 60.0% |
| **Baseline Laboratory Tests** | | |
| CRP (mg/L) | 8.93 (4.30, 27.17) | |
| ESR (mm/h) | 39 (27, 50) | |
| eGFR(mL/min) | 74.63 (24.87, 102.76) | |
| Scr (μmol/L) | 87 (60, 237) | |
| IgG4 (g/L) | 0.46 (0.41, 1.04) | |
| IgG (g/L) | 15.61 (12.25, 17.16) | |
| IgM (g/L) | 0.80 (0.43, 1.06) | |
| IgE (IU/mL) | 37 (8, 64) | |
| IgA (g/L) | 2.36 (1.79, 3.76) | |
| Hemoglobin (g/L) | 118 (104, 134) | |
| Thickness of RPF Mass (mm) | 29 (23, 31) | |
| Craniocaudal RPF Length (mm) | 93 (75, 114) | |
| **Pathological features (n=5)** | | |
| <10 $IgG4^+$ plasma cells/HPF* | 1 | 20.0% |
| 10-50 $IgG4^+$ plasma cells/HPF | 2 | 40.0% |
| >50 $IgG4^+$ plasma cells/HPF | 2 | 40.0% |
| $IgG4^+/IgG^+$ ≤40% | 4 | 80.0% |
| $IgG4^+/IgG^+$ >40% | 1 | 20.0% |

* HPF: high power field microscopy. Here refers to the number of $IgG4^+$ plasma cells under the high power field microscopy.

**Supplementary Table 3. Labs test results in summary.** Time points refer to prior to the treatment (baseline), 12 weeks of treatment (12-week), and 48 weeks of treatment (48-week).

| Lab Test | Unit | Normal Range | Time Points | Mean | Standard Deviation | 25 percentile | 50 percentile | 75 percentile |
|---|---|---|---|---|---|---|---|---|
| Fibrous Tissue Axial Diameter | mm | - | baseline | 89 | 28 | 75 | 93 | 114 |
| | | | 12-week | 59 | 45 | 9 | 68 | 98 |
| | | | 48-week | 54 | 47 | 0 | 70 | 95 |
| Fibrous Tissue Thickness | mm | - | baseline | 28 | 7 | 23 | 29 | 31 |
| | | | 12-week | 15 | 10 | 4 | 18 | 23 |
| | | | 48-week | 12 | 11 | 0 | 16 | 21 |
| C-Responsive Protein (CRP) | mg/L | ＜3.00 | baseline | 16.02 | 19.24 | 4.30 | 8.93 | 27.17 |
| | | | 12-week | 7.43 | 13.54 | 0.43 | 1.48 | 10.43 |
| | | | 48-week | 7.03 | 12.13 | 0.80 | 1.26 | 9.16 |
| erythrocyte sedimentation rate (ESR) | mm/h | 0-20 | baseline | 39 | 12 | 27 | 39 | 50 |
| | | | 12-week | 17 | 21 | 4 | 9 | 19 |
| | | | 48-week | 21 | 23 | 9 | 12 | 26 |
| serum creatinine (SCR) | μmol/L | 45-84 (women); 59-104 (men) | baseline | 135 | 110 | 60 | 87 | 237 |
| | | | 12-week | 90 | 43 | 62 | 82 | 96 |
| | | | 48-week | 88 | 54 | 59 | 73 | 83 |
| estimated glomerular filtration rate (eGFR) | mL/min | 80.00 - 120.00 | baseline | 66.58 | 37.10 | 24.87 | 74.63 | 102.76 |
| | | | 12-week | 76.87 | 25.41 | 64.79 | 77.44 | 98.34 |
| | | | 48-week | 82.21 | 26.55 | 73.54 | 87.84 | 100.57 |
| Hemoglobin | g/L | 115-150 | baseline | 116 | 19 | 104 | 118 | 134 |
| | | | 12-week | 133 | 21 | 118 | 131 | 145 |
| | | | 48-week | 130 | 14 | 118 | 132 | 143 |
| IgG4 | g/L | 0.03-2.01 | baseline | 0.70 | 0.43 | 0.41 | 0.46 | 1.04 |
| | | | 12-week | 0.22 | 0.10 | 0.15 | 0.23 | 0.30 |
| | | | 48-week | 0.47 | 0.43 | 0.22 | 0.27 | 0.90 |
| IgG | g/L | 7.00 - 16.00 | baseline | 15.07 | 2.70 | 12.25 | 15.61 | 17.16 |
| | | | 12-week | 9.65 | 1.54 | 8.20 | 9.30 | 11.18 |
| | | | 48-week | 9.47 | 4.59 | 8.66 | 9.20 | 11.58 |
| IgE | IU/mL | ＜100 | baseline | 36 | 26 | 8 | 37 | 64 |

| | | | | | | | | |
|---|---|---|---|---|---|---|---|---|
| | | | 12-week | 20 | 13 | 13 | 18 | 29 |
| | | | 48-week | 39 | 82 | 5 | 9 | 15 |
| IgM | g/L | 0.40 - 2.30 | baseline | 0.80 | 0.35 | 0.43 | 0.80 | 1.06 |
| | | | 12-week | 0.81 | 0.41 | 0.47 | 0.74 | 1.09 |
| | | | 48-week | 0.87 | 0.31 | 0.52 | 0.80 | 1.10 |
| IgA | g/L | 0.70 - 4.00 | baseline | 2.77 | 1.38 | 1.79 | 2.36 | 3.76 |
| | | | 12-week | 2.11 | 1.26 | 1.40 | 1.58 | 2.59 |
| | | | 48-week | 2.09 | 0.89 | 1.33 | 2.15 | 2.82 |

**Supplementary Table 4**. Antibodies used in flow cytometric analysis.

| **Antibodies** | **Source** | **Identifier** |
|---|---|---|
| Alexa Fluor® 700 anti-human CD3 Antibody | Biolegend | Cat:317340; RRID:AB_2563408 |
| FITC anti-human CD4 Antibody | Biolegend | Cat:357406; RRID: AB_2562357 |
| PerCP anti-human CD8 Antibody | Biolegend | Cat:344708; RRID: AB_1967149 |
| PE anti-human CD25 Antibody | Biolegend | Cat:302606; RRID: AB_314276 |
| Brilliant Violet 510™ anti-human CD45RA Antibody | Biolegend | Cat:304142; RRID: AB_2561947 |
| Brilliant Violet 605™ anti-human CD127 (IL-7Rα) Antibody | Biolegend | Cat:351334; RRID: AB_2562022 |
| Brilliant Violet 421™ anti-human CD197 (CCR7) Antibody | Biolegend | Cat:353208; RRID: AB_11203894 |
| Brilliant Violet 711™ anti-human TCR Vα7.2 Antibody | Biolegend | Cat:351732; RRID: AB_2629680 |
| PE/Dazzle™ 594 anti-human CD161 Antibody | Biolegend | Cat:339940; RRID: AB_2565868 |
| Brilliant Violet 650™ anti-human CD196 (CCR6) Antibody | Biolegend | Cat:353426; RRID: AB_2563869 |
| Alexa Fluor® 647 Rat Anti-Human CXCR5 (CD185) | BD Biosciences | Cat:558113; RRID: AB_2737606 |
| CD279 (PD-1) Monoclonal Antibody (eBioJ105 (J105)), Biotin, eBioscience™ | Thermo Fisher | Cat:13-2799-82; RRID: AB_837120 |
| CD194 (CCR4) Monoclonal Antibody (D8SEE), APC, eBioscience™ | Thermo Fisher | Cat:17-1949-42; RRID: AB_2573176 |
| PE-Cy™7 Streptavidin (SA) | BD Biosciences | Cat:557598; RRID: AB_10049577 |
| PE-CF594 Mouse Anti-Human CD183 (CXCR3) | BD Biosciences | Cat:562451; RRID: AB_11153118 |
